# Potential Structural Biomarkers in 3D Images Validated by the First Functional Biomarker for Early Age-Related Macular Degeneration - ALSTAR2 Baseline

**DOI:** 10.1101/2023.09.10.23295309

**Authors:** Sohaib Fasih-Ahmad, Ziyuan Wang, Zubin Mishra, Charles Vatanatham, Mark E Clark, Thomas A. Swain, Christine A. Curcio, Cynthia Owsley, SriniVas R Sadda, Zhihong Jewel Hu

## Abstract

**Purpose:** While intermediate and late age-Related Macular Degeneration (AMD) have been widely investigated, rare studies were focused on the pathophysiologic mechanism of early AMD. Delayed rod-mediated dark adaptation (RMDA) is the first functional biomarker for incident early AMD. The status of outer retinal bands on optical coherence tomography (OCT) may be potential imaging biomarkers and the purpose is to investigate the hypothesis that the integrity of interdigitation zone (IZ) may provide insight into the health of photoreceptors and retinal pigment epithelium (RPE) in early AMD.

**Methods:** We establish the structure-function relationship between ellipsoid zone (EZ) integrity and RMDA, and IZ integrity and RMDA in a large-scale OCT dataset from eyes with normal aging (n=237), early AMD (n=138), and intermediate AMD (n=101) by utilizing a novel deep-learning-derived algorithm with manual correction when needed to segment the EZ and IZ on OCT B-scans (57,596 B-scans), and utilizing the AdaptDx device to measure RMDA.

**Results:** Our data demonstrates that slower RMDA is associated with less preserved EZ (r = -0.334; p<0.001) and IZ area (r = -0.591; p<0.001), and decreased IZ thickness (r = -0.434; p<0.001). These associations are not apparent when considering normal eyes alone.

**Conclusions:** The association with IZ area and RMDA in large-scale data is biologically plausible because retinoid availability and transfer at the interface attributed to IZ is rate-limiting for RMDA. This study supports the hypothesis that the IZ integrity provides insight into the health of photoreceptors and RPE in early AMD and is a potential new imaging biomarker.

## Introduction

Age-related macular degeneration (AMD) is a leading cause of irreversible central vision impairment worldwide^1^ and subsequently causes significant psychological and socioeconomic burden due to depression, anxiety, and difficulty reading and driving.^2-4^ Currently, strategies to reduce this burden focus on preventing or stabilizing end-stage neovascular and atrophic processes.^5^ However, the vast majority of individuals with AMD have early disease^6^ and there are no proven means to arrest the progression of early AMD, nor prevention strategies for those at high risk. A key barrier to developing preventative and therapeutic strategies is the lack of valid and responsive endpoints.^7^

Optical coherence tomography (OCT) is currently used to visualize 3D changes in retinal structure. In AMD, OCT biomarkers are used for diagnosis, management and for monitoring progression of the disease.^8, 9^ This study, concentrates on two hyperreflective bands seen on OCT termed the ellipsoid zone (EZ) and interdigitation zone (IZ) based on current consensus nomenclature.^10, 11^ OCT segmentation can be a time-consuming and tedious process.^12^ Therefore, we applied a novel, deep-learning-derived, graph-based algorithm^13^ for the automated segmentation that was manually corrected when needed to adequately analyze a large sample of B-scans in 3D OCT in an efficient yet highly accurate manner.

For regulatory approval, novel imaging biomarkers should have biologic plausibility by providing a biologic, physiologic, or pathologic pathway for the association of the biomarker with the disease and should have a contextual linkage between a biomarker and its intended use.^14^ The current understanding of the hyperreflective EZ signal in spectral domain OCT is that it is dominated by mitochondria in photoreceptor inner segments,^15-17^ and mitochondrial dysfunction contributes to the outer retinal degeneration seen in AMD.^18-20^ Further, the hyperreflective IZ signal is attributed to the interface of photoreceptor outer segments, apical processes of the retinal pigment epithelium (RPE) and the surrounding interphotoreceptor matrix.^21^ The RPE supports the photoreceptors at this interface by transferring substances locally produced by the RPE or substances ultimately derived from circulation, such as retinoids, that are required for phototransduction and photoreceptor health.^22^

Delayed rod-mediated dark adaptation (RMDA) is a slower return to rod-mediated sensitivity following a bright light flash stimulus. RMDA was established as functional biomarker for incipient early AMD by the Alabama Study on Age-related Macular Degeneration (ALSTAR).^23^ The retina has the highest metabolic demand in the body and this metabolic demand is even greater in the dark when ionic currents are flowing into the outer segments.^24-26^ Given this metabolic demand in transitioning to the dark, it is plausible that mitochondrial dysfunction may play a role in delayed RMDA. Additionally, RMDA entails multiple steps which includes retinoid transfer from RPE to photoreceptors, and retinoid availability is a rate-limiting for the speed of RMDA.^27, 28^ Consequently, one might hypothesize that delayed RMDA may be closely associated with changes in the EZ and IZ on OCT. **While The EZ has been shown to correlate with visual acuity, the relationships between EZ integrity and RMDA, as well as IZ integrity and RMDA, have not been well-defined**.

In this study, we utilize automated OCT segmentation with manual correction to explore the potential of EZ and IZ area on OCT as an imaging biomarker for early AMD by measuring their association with delayed RMDA.

## Methods

The Alabama Study on Early Age-Related Macular Degeneration 2 (ALSTAR2) is a prospective cohort study on normal aging and early and intermediate AMD whose purpose is to validate retinal imaging biomarkers in these conditions with visual function measures (Clinicaltrials.gov identifier NCT04112667, October 7, 2019)^29^. The study was approved by the Institutional Review Board of the University of Alabama at Birmingham. All participants provided written informed consent after the nature and purpose of the study were explained. Conduct of the study followed the Declaration of Helsinki. The baseline data from ALSTAR2 were collected between October 2019 and September 2021, which included a 4-month pause in enrollment due to the coronavirus pandemic (March-June 2020).

Participants ≥ 60 years old were recruited from the Callahan Eye Hospital Clinics, the clinical service of the University of Alabama at Birmingham Department of Ophthalmology and Visual Sciences. Three groups were recruited: those with early AMD and intermediate AMD, and those in normal macular health. The clinic’s electronic health record was used to search for patients with early or intermediate AMD using International Classification of Diseases 10 codes (H35.30*; H35.31*; H35.36*). One of the investigators (C.O.) screened charts to confirm that participants met the eligibility criteria. Exclusion criteria were (1) any eye condition or disease in either eye (other than early cataract) in the medical record that can impair vision including diabetic retinopathy, glaucoma, ocular hypertension, history of retinal diseases (e.g., retinal vein occlusion, retinal degeneration), optic neuritis, corneal disease, previous ocular trauma or surgery, and refractive error ≥ 6 diopters; (2) neurological conditions that can impair vision or judgment including multiple sclerosis, Parkinson’s disease, stroke, Alzheimer’s disease, seizure disorders, brain tumor, traumatic brain injury; (3) psychiatric disorders that could impair the ability to follow directions, answer questions about health and functioning, or to provide informed consent; (4) diabetes; (5) any medical condition that causes significant frailty or was thought to be terminal. Persons in normal macular health met the same eligibility criteria except they were not classified with the International Classification of Diseases – 10 (ICD-10) codes indicative of AMD. Letters were sent to potential participants, with the study coordinator following up by phone to determine interest.

One eye was tested for each participant, with the eye selected for testing being the eye with better acuity. If the eyes had the same acuity, then an eye was randomly selected. Classification into the 3 groups was based on a trained grader’s (M.E.C.) evaluation of 3-field color fundus photographs taken with a digital camera (FF-450, Carl Zeiss Meditec) following dilation with 1% tropicamide and 2.5% phenylephrine hydrochloride. The Age-Related Eye Disease Study (AREDS) 9-step classification system^30^ was used by the grader to identify AMD presence and severity. Group membership was determined, as follows: eyes in normal macular health had AREDS grade 1, early AMD had grades 2 to 4, and intermediate AMD had grades 5 to 8. We also used the Beckman classification system^31^ with normal aging defined to include grades 1 to 2, early AMD as grade 3, and intermediate AMD as grade 4. The grader was masked to all other participant characteristics. As previously described,^32^ intra-grader agreement was K = 0.88; intergrader agreement with a second grader was K = 0.75. Demographic information for birthdate, gender, and race/ethnicity were obtained through a self-administered questionnaire.

Rod-mediated dark adaptation was assessed with the AdaptDx device (Lumithera, Poulsbo WA). Testing occurred in a dark, light-tight room after dilation. RMDA was measured on the superior vertical meridian at 5° eccentricity to probe the area of proportionately greatest rod loss in aging and AMD.^33, 34^ The procedure began with a photo-bleach exposure to a 6° diameter flash centered at each test target location (equivalent ∼83% bleach; 50 ms duration, 58,000 scotopic cd/m^2^ s intensity^35^) while the participant focused on the fixation light. Threshold measurement (3-down/1 up threshold strategy) for a 2° diameter, 500 nm circular target began 15 seconds after bleach offset. The participant was instructed to maintain fixation and press a button when the flashing target first became visible. Log thresholds were expressed as sensitivity in decibel units as a function of time since bleach offset. Threshold measurement continued at 30-second intervals until the RIT was reached. Rod intercept time is the duration in minutes required for sensitivity to recover to a criterion value of 5.0 x 10^-3^ scotopic cd/m^2^,^23, 36^ located in the latter half of the second component of rod-mediated recovery.^27, 37^ If RIT was not reached, the threshold measurement procedure stopped at 45 minutes. For some participants where the threshold measurement procedure was stopped, the AdaptDx’s algorithm generated a RIT if it could be computed based on previous thresholds. Participants with fixation errors > 30% were excluded from analysis.

We acquired spectral-domain OCT volumes (Spectralis HRA⍰+ ⍰OCT, Heidelberg Engineering, Heidelberg, Germany; λ⍰= ⍰870⍰nm; scan depth, 1.9⍰mm; axial resolution, 3.5⍰μm per pixel in tissue; lateral resolution, 14⍰μm per pixel in tissue), with Automatic Real-Time averaging >⍰9, and quality (signal-to-noise) 20–47⍰dB. B-scans (n⍰= ⍰121 scans, spacing =60⍰μm) were horizontally oriented and centered over the fovea in a 30°×⍰25° (8.6⍰× ⍰7.2⍰mm) area.

The de-identified raw OCT B-scan volumes were imported into 3D-OCTOR, a previously described and validated SD-OCT reading center grading software.^38-40^ A novel deep-learning-derived graph-based algorithm was applied to segment the inner and outer boundaries of the IZ and the outer boundary of the EZ.^13^ Errors in automated segmentation were manually corrected and the boundaries were deleted if either band (EZ or IZ) was not clearly discernible in a region of scan, by a trained, masked grader (SFA).

For the EZ, the inner border of the hyperreflective EZ band was segmented by the algorithm and then manually adjusted as needed by the grader. The inner border of the EZ was defined as the hyperreflective pixel immediately subjacent to the overlying hyporeflective pixel of the myoid zone. If the grader was unable to distinguish the hyperreflective EZ band from the hyporeflective band of the myoid zone or the unnamed hyporeflective band between the EZ and IZ at any point on the B-scan, the segmentation was deleted. Subsequently, the area of EZ on that portion of the B-scan and half the distance to each adjacent B-scan was considered not discernible or not preserved.

Similarly, for the IZ, both the inner and outer boundaries were segmented by the algorithm and manually adjusted by the grader. The inner boundary of the IZ was defined as the hyperreflective pixel subjacent to the overlying hyporeflective band between the EZ and IZ. The outer boundary was defined as the hyperreflective pixel suprajacent to the underlying hyporeflective band between the IZ and RPE. If the grader was unable to distinguish between the IZ and either the underlying or overlying hyporeflective bands the inner boundary was merged with the outer boundary. This results in the area of IZ on that portion of B-scan and half the distance to each adjacent B-scan to be considered not discernible (or not preserved), with a thickness of zero.

Figure 1 demonstrates examples of EZ and IZ grading in no AMD, early AMD, and intermediate AMD eyes. Figure 2 shows further examples of EZ and IZ grading in normal, early AMD, and intermediate AMD eyes on B-scans that include the fovea.

**Figure 1.**
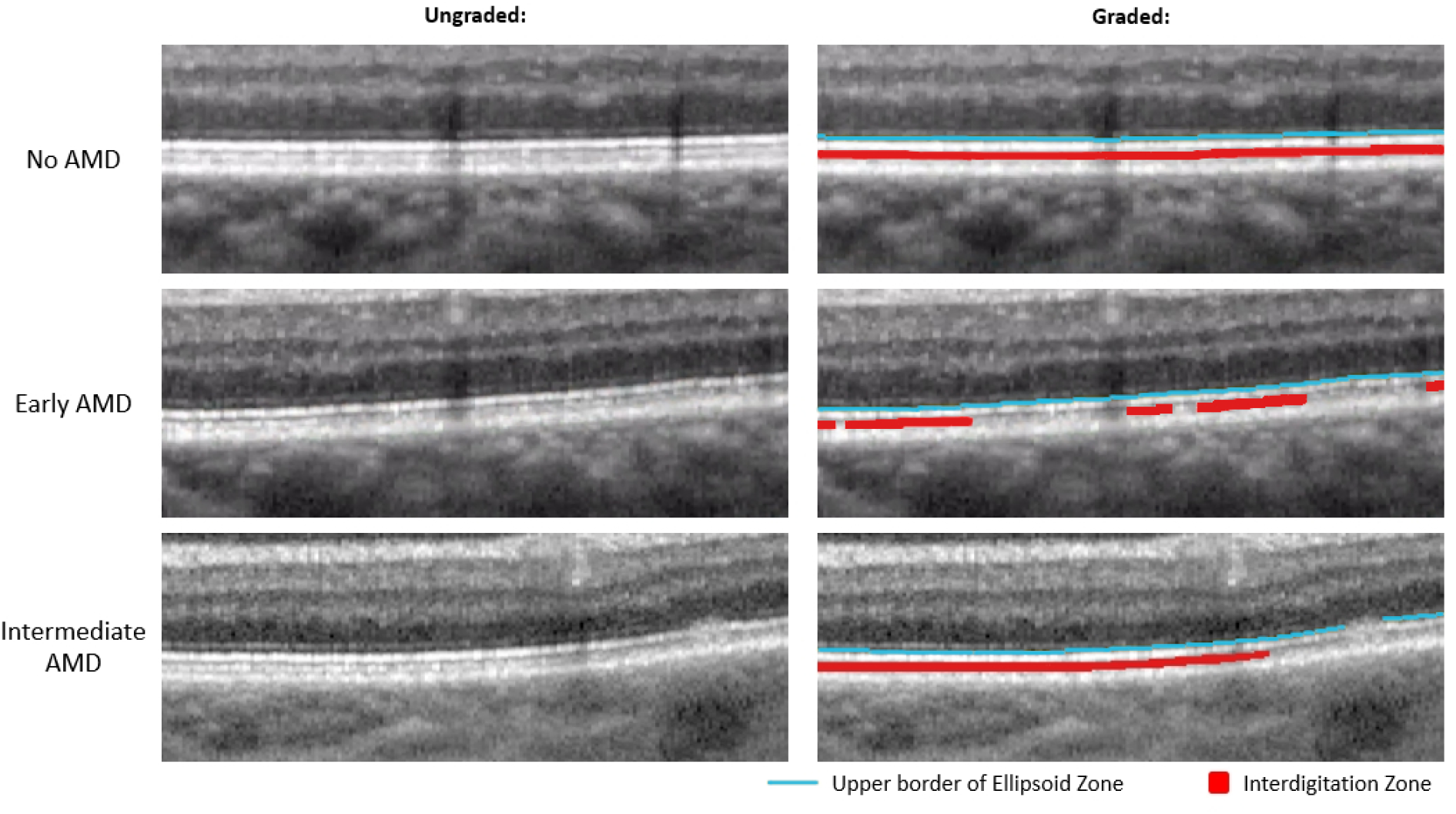
Magnified view of outer retinal bands in no AMD, early AMD, and intermediate AMD eyes. *Left:* Unaltered magnified OCT B-scans. *Right*: Same magnified OCT B-scan as the left side but graded for EZ and IZ. Yellow line is the upper border of the EZ which was used as a proxy for discernable EZ, Red area represents discernable IZ.

**Figure 2.**
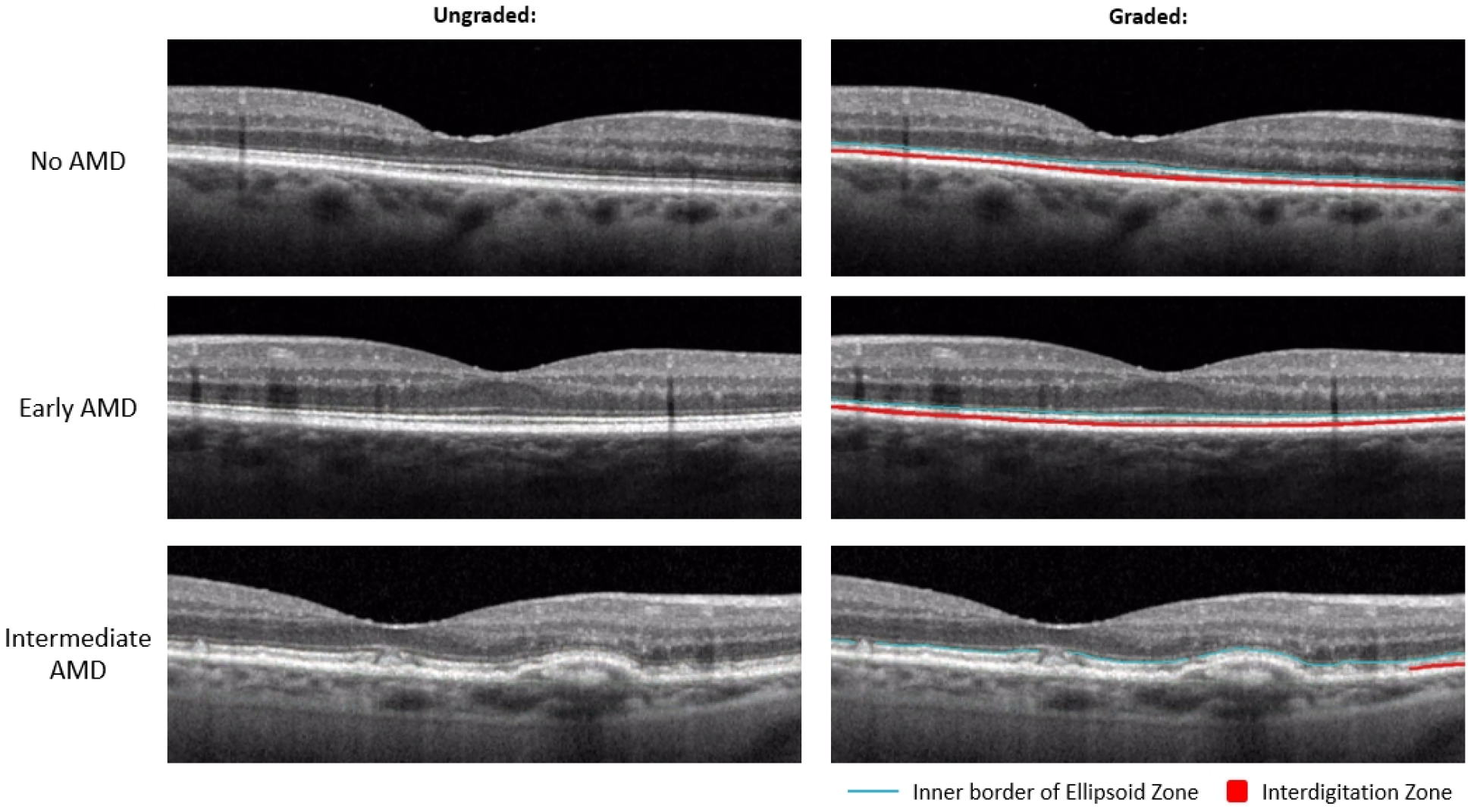
Magnified view of outer retinal bands on foveal B-scans in no AMD, early AMD, and intermediate AMD eyes. *Left*: Unaltered magnified OCT B-scans. *Right:* Same magnified OCT B-scan as the left side but graded for EZ and IZ. Yellow line is the upper border of the EZ which was used as a proxy for discernable EZ, Red area represents discernable IZ.

Mean area of IZ, mean thickness of IZ and mean area of EZ were computed in the entire fovea-centered Early Treatment of Diabetic Retinopathy Study (ETDRS) grading grid,^41^ as well as within the central subfield, inner ring, and outer ring of the ETDRS grid, as implemented in the Spectralis software (radii of 0.5, 1.0-1.5, and 1.5-3.0 mm, respectively) . Separate means were calculated for each of the 3 groups based on AREDS 9-step classification system^30^: normal, early AMD, and intermediate AMD. Differences in the groups were assessed using analysis of covariance (ANCOVA). The relationships between RIT and IZ area, IZ thickness, and EZ area were assessed using Spearman correlation coefficients (r) and were age-adjusted. Spearman correlation coefficients were calculated for the entire ETDRS grid as well as the central, superior inner, and superior outer subfields for each of the 3 groups (normal eyes, early AMD, and intermediate AMD). The level of significance was p ≤0.05 (two-sided).

## Data Availability

The data and code generated during the study is accessible from the corresponding author based on reasonable request and subject to the rule/regulatory of the involved institutes.

## Results

A total of 476 eyes were assessed, of which 237 were classified as no AMD, 138 were classified as early AMD and 101 were classified as intermediate AMD. Demographics of this population are shown in Table 1.

**Table 1.** Demographic Characteristics of Participants (n = 476)

| **Table 1. Demographic Characteristics of Participants (n = 476)** |  |  |
| --- | --- | --- |
| **Variable Names and Groups** | **Mean** | **SD or n%** |
| Age (years) | 71.75 | 5.88 |
| Age group |  |  |
| 60-69 | 166 | 34.87 |
| 70-79 | 265 | 55.67 |
| 80-89 | 45 | 9.45 |
| Gender |  |  |
| Male | 192 | 40.34 |
| Female | 284 | 59.66 |
| Race |  |  |
| White | 431 | 90.55 |
| Black | 40 | 8.40 |
| Other* | 5 | 1.05 |

SD = Standard Deviation
\*Other includes 4 Asians or Pacific Islanders and 3 Native Americans.

The area of EZ on OCT is reduced in eyes with intermediate AMD compared to eyes with early AMD and is reduced in eyes with early AMD compared to eyes without AMD in the entire ETDRS grid as well as each of the subfields that were analyzed. Figure 3 demonstrates the en face maps generated by 3D-OCTOR representing the area of EZ and IZ within the ETDRS grid and the associated subfields.

**Figure 3.**
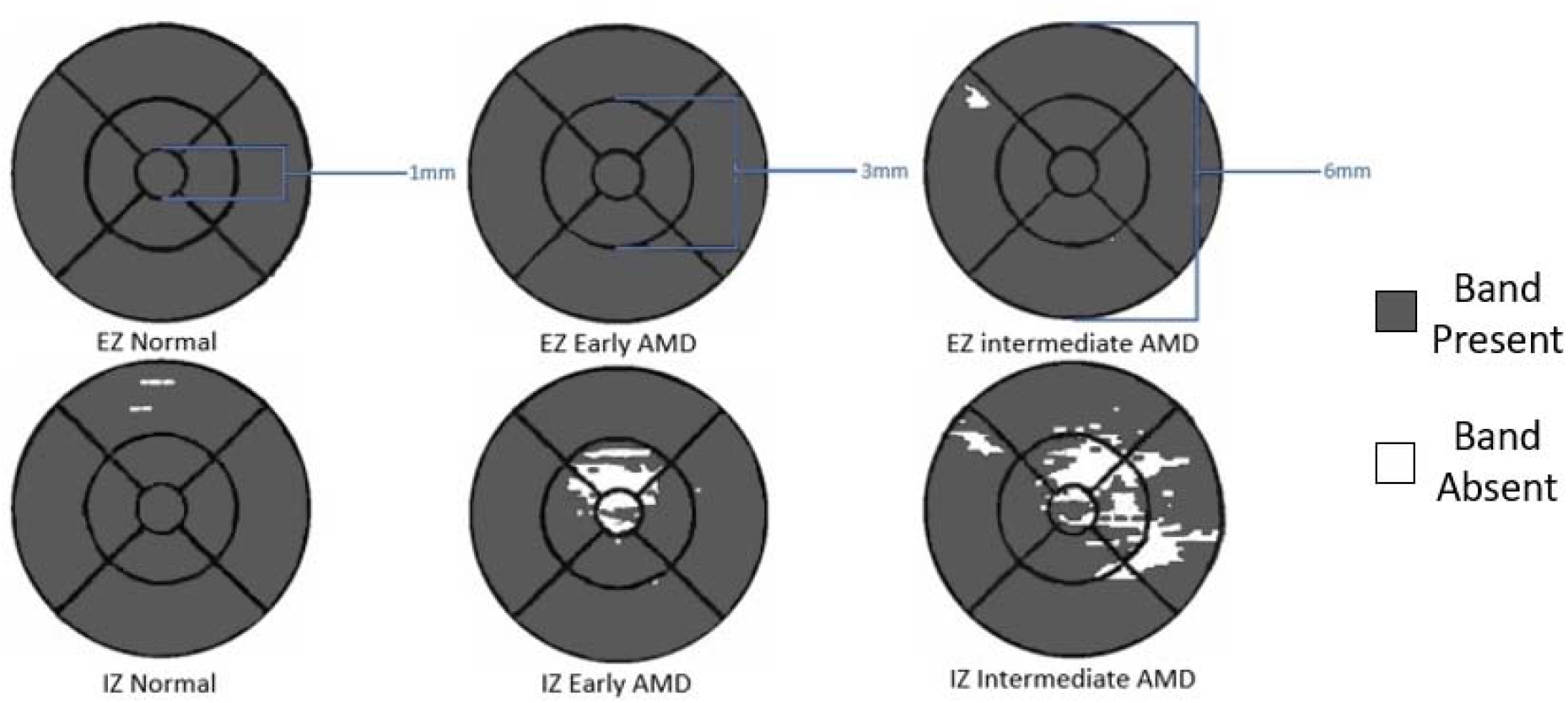
En face maps of the EZ and IZ in a normal aging, early AMD, and intermediate AMD eye within the ETDRS grid. The diameter of each subsection (central subfield, inner ring, outer ring) are labeled in blue. White regions indicate absence of the band (EZ or IZ); gray region indicate areas where the band is present

Slower RMDA (longer RIT) was associated with less preserved EZ area (r= -0.334) in the entire ETDRS grid. The strength of this association was diminished, but still present, when considering each of the AREDS categories separately. Likewise, the same association was observed in each of the ETDRS subfields that were considered, and these associations were weaker than that of the entire ETDRS grid. In the central subfield, no correlation between EZ area and RIT was found when considering eyes in each of the AREDS categories separately. In the inner ring, slower RMDA was associated with less preserved EZ area only in eyes with intermediate AMD. In the outer ring, a similar association was observed with all eyes considered and in each of the AREDS categories considered separately. Table 2A and Figure 4 shows the associations for the various evaluated subfields.

**Table 2A.** Area of preserved EZ/IZ and correlation between RMDA and EZ/IZ areas in the entre grid, central subfield, and inner and outer rings of the ETDRS grid in all eyes and subdivided by AREDS categories (normal, early AMD., intermediate AMD).

| Table 2A |  | Correlation of RMDA at 5 degrees with Area of EZ and IZ |  |  |  |  |  |
| --- | --- | --- | --- | --- | --- | --- | --- |
|  |  |  | Area |  |  | Correlation |  |
| Layer | ETDRS Subfield | AREDS Category | Mean | Std Dev | p-value | Correlation Coefficient (r) | p-value |
| Ellipsoid Zone (EZ) | Entire Grid | All | 28.149 | 0.068 | <.0001 | -0.334 | <.0001 |
|  |  | Normal | 28.270 | 0.023 |  | -0.199 | 0.002 |
|  |  | Early | 28.246 | 0.111 |  | -0.272 | 0.001 |
|  |  | Intermediate | 27.699 | 1.423 |  | -0.268 | 0.007 |
|  | Central Subfield | All | 0.774 | 0.065 | <.0001 | -0.217 | <.0001 |
|  |  | Normal | 0.790 | 0.003 |  | -0.055 | 0.401 |
|  |  | Early | 0.787 | 0.014 |  | -0.039 | 0.652 |
|  |  | Intermediate | 0.752 | 0.088 |  | -0.033 | 0.743 |
|  | Inner Ring | All | 6.232 | 0.281 | <.0001 | -0.348 | <.0001 |
|  |  | Normal | 6.280 | 0.011 |  | -0.021 | 0.746 |
|  |  | Early | 6.273 | 0.041 |  | -0.117 | 0.175 |
|  |  | Intermediate | 6.088 | 0.373 |  | -0.227 | 0.023 |
|  | Outer Ring | All | 21.0893 | 1.058 | <.0001 | -0.292 | <.0001 |
|  |  | Normal | 21.2091 | 0.016 |  | -0.291 | <.0001 |
|  |  | Early | 21.1947 | 0.079 |  | -0.322 | <.0001 |
|  |  | Intermediate | 20.8628 | 1.128 |  | -0.258 | 0.010 |
| Interdigitation Zone (IZ) | Entire Grid | All | 27.125 | 3.219 | <.001 | -0.591 | <.0001 |
|  |  | Normal | 28.250 | 0.136 |  | -0.442 | <.0001 |
|  |  | Early | 27.909 | 0.777 |  | -0.496 | <.0001 |
|  |  | Intermediate | 23.223 | 5.538 |  | -0.472 | <.0001 |
|  | Central Subfield | All | 0.707 | 0.192 | <.001 | -0.531 | <.0001 |
|  |  | Normal | 0.789 | 0.011 |  | -0.069 | 0.288 |
|  |  | Early | 0.766 | 0.065 |  | -0.257 | 0.001 |
|  |  | Intermediate | 0.436 | 0.288 |  | -0.288 | 0.004 |
|  | Inner Ring | All | 5.863 | 1.060 | <.001 | -0.584 | <.0001 |
|  |  | Normal | 6.275 | 0.032 |  | -0.107 | 0.101 |
|  |  | Early | 6.173 | 0.235 |  | -0.303 | <.0001 |
|  |  | Intermediate | 4.397 | 1.646 |  | -0.414 | <.0001 |
|  | Outer Ring | All | 20.555 | 2.190 | <.001 | -0.538 | <.0001 |
|  |  | Normal | 21.195 | 0.122 |  | -0.460 | <.0001 |
|  |  | Early | 20.974 | 0.637 |  | -0.460 | <.0001 |
|  |  | Intermediate | 18.389 | 4.125 |  | -0.450 | <.0001 |

**Table 2B.** Mean thickness of IZ and correlation between RMDA and IZ in the entire grid, central subfield, and inner and outer rings of the ETDRS grid in all eves and subdivided by AREDS categories (normal, early AMD. intermediate AMD).

| Table 2B |  | Correlation of RMDA at 5 degrees with Mean Thickness of IZ |  |  |  |  |  |
| --- | --- | --- | --- | --- | --- | --- | --- |
|  |  |  | Mean Thickness |  |  | Correlation |  |
| Layer | ETDRS Subfield | AREDS Category | Mean | Std Dev | p-value | Correlation Coefficient (r) | p-value |
| Interdigitation Zone (IZ) | Entire Grid | All | 16.221 | 4.093 | <.001 | -0.434 | <.0001 |
|  |  | Normal | 16.983 | 3.572 |  | -0.142 | 0.029 |
|  |  | Early | 17.043 | 3.580 |  | -0.267 | 0.002 |
|  |  | Intermediate | 13.396 | 4.848 |  | -0.508 | <.0001 |
|  | Central Subfield | All | 16.295 | 5.947 | <.001 | -0.482 | <.0001 |
|  |  | Normal | 18.245 | 3.723 |  | -0.082 | 0.210 |
|  |  | Early | 18.225 | 4.428 |  | -0.257 | 0.002 |
|  |  | Intermediate | 9.505 | 7.174 |  | -0.288 | 0.004 |
|  | Inner Ring | All | 15.734 | 4.684 | <.001 | -0.474 | <.0001 |
|  |  | Normal | 16.987 | 3.731 |  | -0.144 | 0.027 |
|  |  | Early | 17.000 | 3.746 |  | -0.231 | 0.007 |
|  |  | Intermediate | 11.257 | 5.480 |  | -0.431 | <.0001 |
|  | Outer Ring | All | 16.362 | 4.101 | <.001 | -0.386 | <.0001 |
|  |  | Normal | 16.987 | 3.731 |  | -0.126 | 0.053 |
|  |  | Early | 17.043 | 3.598 |  | -0.268 | 0.002 |
|  |  | Intermediate | 14.238 | 5.091 |  | -0.494 | <.0001 |

**Figure 4A.**
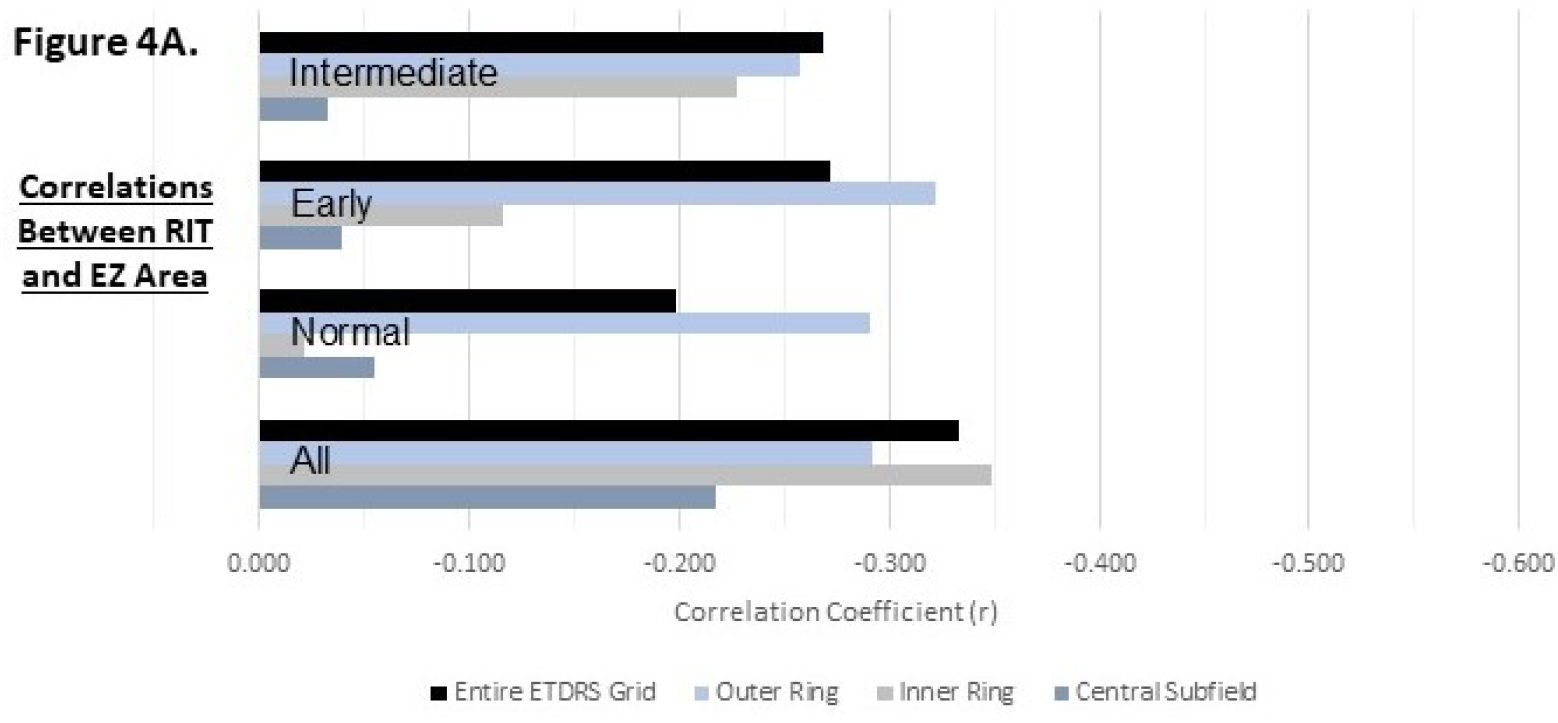
Correlations between RIT and EZ area in the entire ETDRS grid, outer and inner rings, and central subfield of the ETDRS grid for all eyes, and only normaI, early AMD and intermediate AMD eyes

**Figure 4B.**
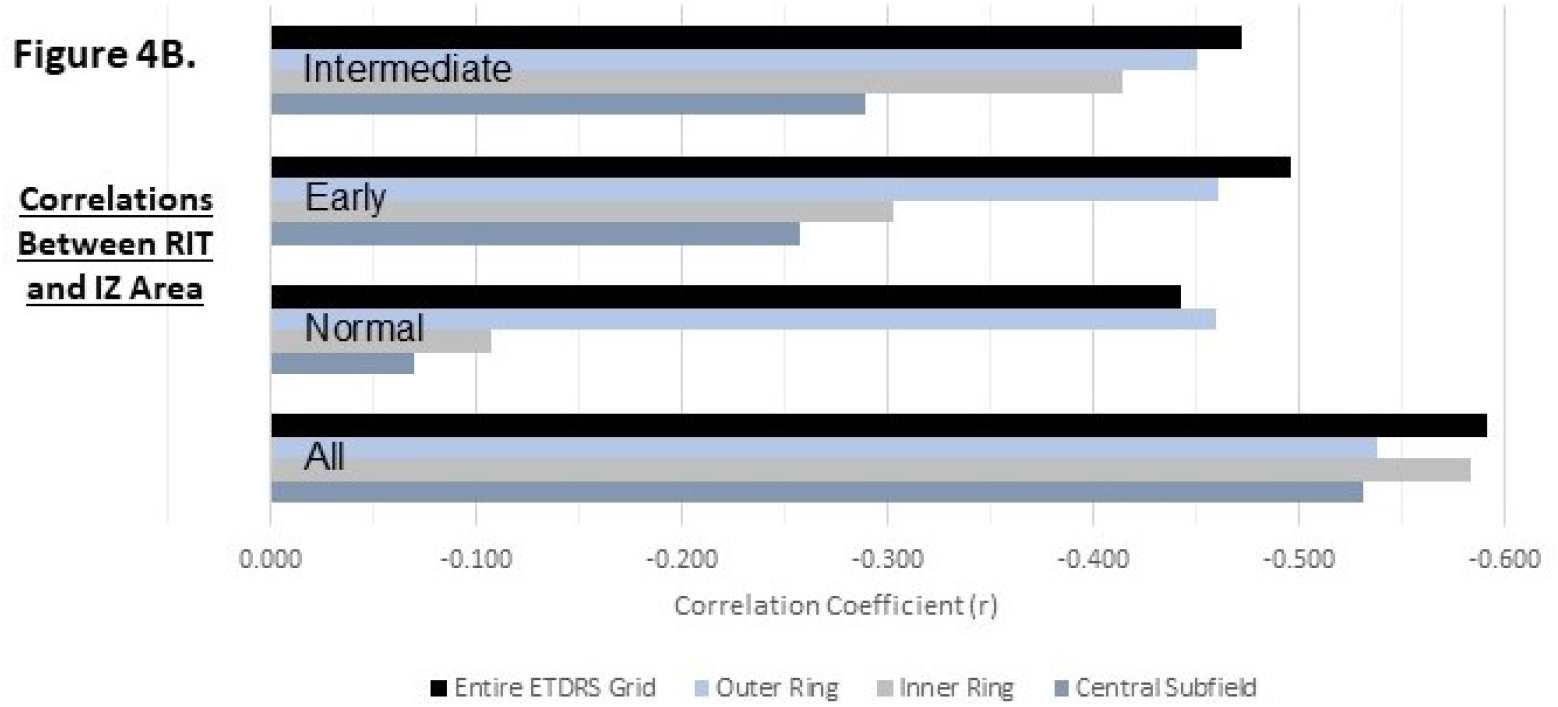
Correlations between RIT and IZ area in the entire ETDRS grid, outer and inner rings, and central subfield of the ETDRS grid for all eyes, and only normaI, early AMD and intermediate AMD eyes

**Figure 4.**
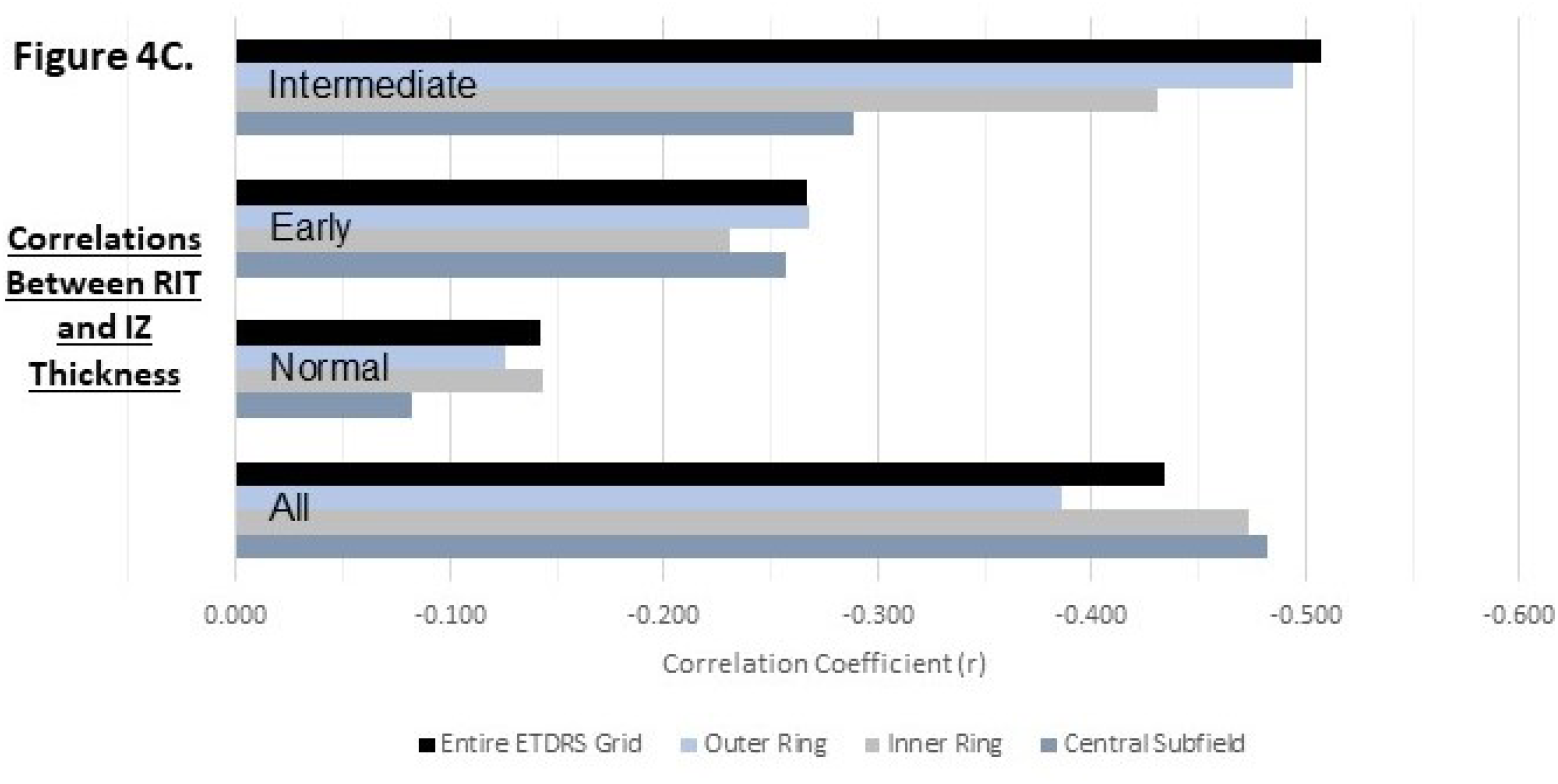
Correlations between RIT and mean IZ thickness in the entire ETDRS grid, outer and inner rings, and central subfield of the ETDRS grid for all eyes, and only normal, early AMD and intermediate AMD eyes

Similar to the EZ, the area of IZ on OCT is decreased in eyes with intermediate AMD compared to eyes with early AMD and is decreased in eyes with early AMD compared to eyes without AMD in the entire ETDRS grid as well as each of the analyzed subfields.

Also, slower RMDA was associated with reduced IZ area for the entire ETDRS grid as well (r= -0.591) and this association was stronger than that between EZ and RIT. This association was again weaker when each of the AREDS categories was considered separately but still notable with correlations of r = -0.442 in normal eyes, r = -0.496 in early AMD eyes, and r = -0.472 in intermediate AMD eyes.

In the central subfield and inner ring, there was no association between IZ area and RIT in normal eyes. There was an association between slower RMDA and IZ area in early AMD and intermediate AMD eyes, but these correlations were notably weaker than the association when considering the entire ETDRS grid.

Conversely, in the outer ring, the association between slower RMDA and IZ area was observed in normal eyes (r = -0.460) and this association was very similar in early AMD eyes (r = -0.460) and intermediate AMD eyes (r = -0.450). Additionally, this association was stronger in the outer ring than in the inner ring or central subfield when each AREDS category was considered separately, but weaker than the association observed while considering the entire grid except in normal eyes.

Associations between slower RMDA and mean thickness of the IZ were also observed for the entire ETDRS grid (r = -0.434) as well as for each of the subfields we analyzed (Table 2B). Notably, these associations were weaker than those between IZ area and RIT. As for the EZ and IZ areas, the associations between RIT and IZ thickness were stronger in eyes with intermediate AMD compared to eyes with early AMD and stronger in eyes with early AMD compared to eyes without AMD. Furthermore, the association between IZ thickness and RIT was strongest in the central subfield, followed by the inner ring and then the outer ring.

## Discussion

In this study, we explored the structure-function relationship between preserved area of the EZ and IZ and mean thickness of the IZ on OCT B-scans with delayed RMDA. We found an association between delayed RMDA and each of these imaging biomarkers with the area of discernible IZ demonstrating the strongest association. This association was also found in the cone-dominant central subfield.

Previously, A.Y. Lee et al using a deep learning model demonstrated that delayed RMDA on foveal OCT B-scans correlates with variable reflectivity of the hyporeflective bands superficial and deep to the EZ.^42^ This variable reflectivity results in a blurring effect that could reduce the discernability of both the EZ and the IZ on OCT, a specific feature that we assessed in this study. The model utilized by Lee et al demonstrated a stronger correlation (r = 0.69) between variable reflectivity of these hyporeflective bands and delayed RMDA^42^ than any of the correlations we calculated between delayed RMDA and discernible EZ or IZ area. However, the strength of the correlation between IZ area in the entire ETDRS grid and RMDA from this study (r = 0.591) were not very dissimilar to this previous study. Conversely, correlation with EZ area was notably weaker (r = 0.334).

The Lee et al study was limited by evaluating only a single B-scan through the fovea while RIT is measured at 5° eccentricity^42^ which may explain the discrepancy between that study and this one. This is because pathologic changes in AMD, such as changes that precede drusen formation, occur most prominently under the fovea.^43-46^ However, pathology under the cone-rich fovea can impact rods in the nearby perifovea where RIT was measured.^47^ As a result, structural changes in the outer retina seen at the fovea may be associated with delayed RMDA. However, our analysis showed slightly weaker correlations in the central foveal subfield of the ETDRS grid than the entire ETDRS grid between the retinal structures we analyzed and RMDA.

In AMD eyes, histopathologic studies have demonstrated preferential loss of rod cells in the macula of eyes with early disease compared with age-matched controls. The greatest loss occurred in the region 0.5-3 mm from the fovea.^34^ This region aligns with the inner ring of the ETDRS grid, where the structure-function association between RMDA and IZ area was strongest with all eyes considered. However, when considering each AREDS category separately, we found the structure-function association was strongest in the outer ring, which contains the RMDA test point (5° eccentricity on the superior vertical meridian), rather than the inner ring or central subfield. This suggests that the observed structure-function association is related to local activity in the outer retina rather than simply a coincident delay in RMDA and decrease in discernible IZ across the entire macula. Additionally, in eyes without AMD, this association was seen in the outer ring only and not in the inner ring or central subfield, suggesting that the effect of local changes may be independent of AMD progression.

Therefore, it is plausible that delayed RMDA results from either central macular pathology, local pathology near the RMDA testing location, or a combination of both. Laíns et al reported that the presence of any abnormalities (EZ disruption, sub-RPE drusen, subretinal drusenoid deposits (SDD), hyperreflective foci, retinal atrophy, subretinal and intraretinal fluid, fibrosis, choroidal neovascularization, and serous pigment epithelial detachment) within the RMDA test spot (located at the same place as ours), as well as any abnormalities in the macula, were significantly associated with longer RIT and therefore delayed RMDA. This association was stronger with abnormalities at the RMDA test spot^48^ which corresponds to our finding that in normal and early AMD eyes in which abnormalities in the entire macula are relatively rare, the structure-function relationship between RMDA and IZ and EZ area are stronger in the ETDRS ring containing the RMDA testing spot as those abnormalities likely contribute to a greater proportion of the loss in EZ and IZ than in intermediate eyes.

SDD, also termed reticular pseudodrusen, first appear within the outer subfields of the ETDRS grid and commonly spread toward the fovea.^49^ Several studies demonstrate that the presence of SDD is associated with markedly delayed RMDA.^48, 50-52^ Stage 1 SDD are defined by diffusely distributed granular hyperreflective material between the RPE and the EZ, blurring the boundaries between normally well delineated hyperreflective IZ and RPE lines. Stage 2 and 3 represent the expansion of hyperreflective material accumulation between the RPE and EZ that further distorts band boundaries, with stage 3 SDD protruding upward though the EZ.^53-55^ SDD abundance in the ETDRS outer ring and progression impacting EZ discernability possibly explains why associations between RMDA and EZ area are stronger in the outer ring of early AMD eyes than in intermediate AMD.

We speculate that the same mechanisms may be responsible for both the decrease in IZ discernability and delay in RMDA. RPE outer segments and apical processes containing melanosomes contribute reflectivity to the IZ^56^ and these RPE apical processes also contain many proteins relevant to the retinoid processing pathways.^57^ The RPE supports the photoreceptors at the IZ by transferring retinoids required for phototransduction^22^ and the availability of retinoids is ultimately rate-limiting for RMDA.^27^ Therefore, a reduction of IZ anatomical substrates could delay RMDA. Furthermore, the RPE is thought to undergo several structural changes with aging, including loss of melanin.^58^ It should be noted that we age-adjusted the correlations to better isolate the impact of AMD on the IZ and RMDA and eliminate the potentially confounding effect of normal aging.

Strengths of this study include a large sample (n = 476) of eyes in a cohort designed to cross-sectionally probe the transition from normal aging to AMD, and a comprehensive 3D OCT analysis of the bands of interest throughout the macula utilizing dense volume scanning (121 B-scans in an 8.6⍰×⍰7.2⍰mm area). While a few of retinal layer segmentation algorithms performed overall well in OCT, ^59-61^ our deep-learning-derived graph-based algorithm^13^ is the only one which can segment all the retinal deposits, and can reflect the subtle IZ changes in early AMD. It allows us to improve our understanding of the functional RMDA by assessing how structural changes in different regions of the macula are associated with RMDA. Limitations include a relatively homogenous population dominated by participants of European descent and an OCT analysis restricted to ETDRS regions rather than the exact RMDA test location. On the other hand, it is unclear how wide an area surrounding the test spot may contribute to the measured RMDA.

In conclusion, **RMDA is correlated with the status of outer retinal bands in early and intermediate AMD eyes. The correlation of IZ with RMDA is stronger than that of EZ**. IZ integrity on OCT fulfills the conditions of a valuable imaging biomarker for early AMD by establishing a structure-function association with delayed RMDA. This relationship is biologically plausible, because retinoid availability and transfer at this interface is rate-limiting for RMDA. The structure-function association of EZ area and RMDA was weaker and likely useful only in later stages of AMD. Future studies assessing other aspects of retinoid availability to the photoreceptors and their relationship to IZ area may offer further support for IZ on OCT as an imaging biomarker for early AMD.

## Acknowledgements

This work was supported by the National Eye Institute of the National Institutes of Health under Award Numbers R01EY029595 and R21EY030619, Research to Prevent Blindness, EyeSight Foundation of Alabama, and the Dorsett Davis Discovery Fund.

